# Worldwide trends in COVID-19 related attacks against healthcare

**DOI:** 10.1101/2023.07.18.23292819

**Authors:** Willeke Duffhues, Dennis Barten, Harald De Cauwer, Luc Mortelmans, Frits van Osch, Derrick Tin, Marion P.G. Koopmans, Gregory Ciottone

## Abstract

**Background:** During the COVID-19 pandemic, violence targeting healthcare reportedly increased. Attacks against healthcare have the potential to impair the public health response and threaten the availability of healthcare services. However, there is little systematic understanding of the extent and characteristics of healthcare attacks in the setting of a pandemic. This study aimed to investigate global trends regarding COVID-19 related attacks against healthcare from January 2020 until January 2023.

**Methodology:** COVID-19 related incidents that occurred between January 2020 and January 2023 were extracted from the Safeguarding Health in Conflict Coalition database and screened for eligibility. Data collected per incident included temporal factors; country; setting; attack and weapon type; perpetrator; motive; number of healthcare workers (HCWs) and patients killed, injured or kidnapped; and whether the incident caused damage to a health facility.

**Results:** This study identified 255 COVID-19 related attacks against healthcare. The attacks occurred globally and throughout the course of the pandemic. Incidents were heterogeneous with regards to motives, attack types and outcomes. At least 18 HCWs were killed, 147 HCWs were injured and 86 facilities were damaged or destroyed. There were two periods with a peak incidence of reports. The first peak occurred during the beginning of the pandemic, and predominantly concerned stigma-related attacks against healthcare. The second peak, in 2021, was mainly composed of conflict-related attacks in Myanmar, and attacks targeting the global vaccination campaign.

**Conclusion:** COVID-19 related attacks against healthcare occurred globally and in a variety of settings throughout the course of the pandemic. The findings of this study can be used to prevent and mitigate healthcare attacks during the ongoing and future pandemics.

## Introduction

Since the onset of the COVID-19 pandemic, health systems have sustained unprecedented pressures. Healthcare workers (HCWs) faced difficult working conditions, including prolonged shifts with intensive use of personal protective equipment, psychological distress and occupational risk of contracting COVID-19. (1) Nearly half of the HCWs experienced burnout during the COVID-19 pandemic. (2) Inadequate protection of HCWs was raised as a major concern by Amnesty International in a report published in the first year of the pandemic (1), and the World Health Organization has listed investment in the healthcare workforce as one of the key priorities for the coming decade. (3) Although the public showed their support for and solidarity with HCWs, an ‘infodemic’ of false information and fake news, fueling stigma and grievances, contributed to outbreaks of protests and violence. (4) Direct attacks against HCWs have been reported globally, contributing to the further distress of an already overwhelmed workforce. (1, 5–7) Ensuring the protection and well-being of frontline workers plays a pivotal role in enhancing global public health response - not only to the COVID-19 pandemic, but also to potential future outbreaks.

Workplace violence is common in the healthcare setting. A systematic review published in 2019 found that more than half of HCWs around the world experienced workplace violence annually. (8) Furthermore, HCWs and healthcare facilities have been shown to be vulnerable soft targets for terrorist attacks. (9,10) Over the last two decades, there has been a disproportionate rise of terrorist attacks against hospitals when compared to other target types, highlighting the vulnerability of these key structures. (11) This vulnerability may have been further exacerbated by the COVID-19 pandemic. (9,10) Historically, pandemics and attacks against healthcare have always coexisted. Reported factors that are thought to contribute to such attacks include fear of the disease, conspiracy theories and anti-government motives. (12) Moreover, religious, socio-cultural and ethnic disputes are aggravated, exacerbating civil unrest. (12) Indeed, during the pandemic year 2020, COVID-19 was found to be a motive for 165 incidents reported to the Global Terrorism Database (GTD), including arson attacks against 5G telephone masts, and attacks against aid workers, hospitals, testing centers and quarantine facilities.(13)

Internationally, efforts have been made to bring attention to attacks against healthcare. (14) However, as set forth by human rights lawyer Leonard Rubenstein, there has been a long period of inaction to protect HCWs. (15) In 2012, the World Health Organization (WHO) adopted the World Health Assembly Resolution 65.20. This resolution aimed to recognize the harm of attacks against healthcare. (16) Since then, research in the field has expanded.

Several international organizations maintain databases recording attacks against healthcare. The International Committee of the Red Cross (ICRC) reported in August 2020 that they had identified 611 incidents of violence, harassment or stigmatization in relation to COVID-19 cases across more than forty countries. (17) The ICRC strictly maintains a neutral attitude to be able to work in conflict environments. For this reason, they do not make their data or methodology public, nor do they attribute attacks to specific perpetrator groups. (15, 18) The WHO also monitors attacks in its Surveillance System for Attacks against Healthcare (SSA) database. The WHO is transparent about the reliability of its data; attacks reported in the SSA database are categorized according to level of certainty. However, the WHO does not keep account of whether attacks are COVID-19 related, nor provides detailed information about the attacks as it is limited by its political position. (15,18)

In 2014, a number of non-governmental organizations (NGOs) formed the Safeguarding Health in Conflict Coalition (SHCC). The SHCC maintains a mixed-methodology public database and brings out reports, mostly focusing on attacks in conflict regions. (19) In contrast to the databases of the ICRC and WHO, the SHCC database is publicly available, contains detailed information and provides a transparent methodology. (18) Since the beginning of the COVID-19 pandemic, the coalition has also been documenting testimonies of COVID-19 related attacks against healthcare, providing opportunities to study the etiology and impact of such attacks. (20)

Attacks against HCWs, both terrorism-related or not, can severely hamper the public health response in crisis situations and there remains a lack of studies investigating this issue. The objective of this study is to investigate trends regarding COVID-19 related attacks against healthcare through analysis of incidents from the SHCC database from 2019 to 2022.

## Methods

### Database

The SHCC is a collaboration of non-governmental organizations (NGOs), health professional organizations and Johns Hopkins University that was launched to raise awareness about attacks on healthcare, strengthen documentation about these attacks and increase accountability of the perpetrators. (21) SHCC member NGO “Insecurity Insight” manages the database for the coalition (19) and updates it bimonthly. Incident data are collected from multiple sources, including media sources, partner agencies, and publicly available databases, for example the WHO’s Surveillance System for Attacks on Healthcare (SSA) database and the Armed Conflict Location & Event Data Project (ACLED) database. More detailed information regarding data collection can be found in the SHCC methodology codebook. (22) The datasets are compiled using the principles of event-based coding. The coding is based on reported information. The organization provides its information including incident description in an interactive online map and as a database in the Humanitarian Data Exchange. (20, 23)

### Definitions

The SHCC database uses the WHO definition of an attack on healthcare: “any act of verbal or physical violence, threat of violence or other psychological violence, or obstruction that interferes with the availability, access and delivery of curative and/or preventive health services. (22)” The SHCC frequently refers to these attacks as ‘incidents’, as the term ‘attack’ is often interpreted to convey intent, although this is not necessary to meet the WHO definition.(24) In this study both terms are used interchangeably. A healthcare worker (HCW) is defined as: “any person working in a professional or voluntary capacity in the provision of health services or who provides direct support to patients, including administrators, ambulance personnel, community health workers, dentists, doctors, government health officials, hospital staff, medical education staff, nurses, midwives, paramedics, physiotherapists, surgeons, vaccination workers, volunteers, or any other health personnel or medics not named here.” (25) A health facility is “any facility that provides direct support to patients, including clinics, hospitals, laboratories, makeshift hospitals, medical education facilities, mobile clinics, pharmacies, warehouses, or any other health facility not named here.” (25) The SHCC regards an incident as COVID-19 related when the incident is “directly linked to COVID-19 health measures or directly interferes with the availability of, access to, and delivery of COVID-related health services. This includes conflict-related incidents when the perpetrator is a conflict party or political incidents when the perpetrator is a state force harming health workers during COVID-19 related protests.” (25) As most perpetrators do not directly communicate their motives for an attack, an incident is categorized as COVID-19 related when the context of the incident suggests such a connection.

### Incident selection

The SHCC includes any incident affecting healthcare workers, facilities, or transport. For this study, incidents that were categorized as COVID-19 related by the SHCC within the time frame December 2019 up to January 2023 were screened for eligibility. Every incident was reviewed manually by researchers WD and DB for inclusion or exclusion based on the incident description. Consensus was reached for all incidents. Duplicate incidents were excluded. Incidents in which a COVID-19 related motive was suggested; attacks on facilities and/or HCWs specifically involved in the treatment or prevention of COVID-19; HCWs being spat at, coughed at, attacked with a liquid or attacked while traveling to/from work; denial of care to COVID-19 patients and attacks involving material dedicated for COVID-19 care were included. Incidents were excluded when they described fraud; arrests of HCWs that did not violate human rights; non-violent protests not obstructing access to healthcare; arrests made at healthcare facilities, governmental healthcare policy; volunteers not working for healthcare organizations; accidents and incidents with an unclear connection to the pandemic or lacking detailed information. In addition to the SHCC database review, SHCC’s newsletters from December 2019 up to January 2023 were screened for any incidents matching the inclusion criteria that were not included in the SHCC database. (26) Incidents describing systematic violence or several attacks were categorized as multiple incidents and included once.

### Data extraction

Data collected per incident included date and year, country, setting, attack type and weapon, perpetrator, assumed motive, number of HCWs killed, injured or kidnapped and whether the attack damaged or destroyed the health facility (see Appendix 1 for categories and definitions). Information regarding setting, attack type, weapon use, perpetrator, motive and affected HCWs, health facilities and ambulances was categorized from the incident description. In case of missing or unclear information, an attempt was made to retrieve the information by examining the original source in which the incident was reported (as indicated in SHCC newsletters) and by conducting a gray literature search using Google (Google LCC.; Mountain View, California USA) in English, German, French and Spanish.

### Data analysis

Collected data were exported into IBM SPSS Statistics 28^th^ edition™ and analyzed descriptively. Chi-squared tests were applied to evaluate the trends of incidents over time and the differences per world region. A p value <05 was considered to be significant.

## Results

### General results

In total, 227 out of 5844 incidents from the SHCC database entered between December 2019 and January 2023 fulfilled the inclusion criteria. Twenty-eight more incidents were identified by the supplementary screening of 57 SHCC newsletters, adding up to 255 incidents in total (Figure 1). Seventeen (6.7%) of the identified incident reports described multiple incidents and in twenty-four incidents (9.4%) the number of HCWs killed or injured was unknown. The incidents occurred in 54 countries and on all continents except Antarctica. A total number of 18 HCWs were killed, 147 were injured, and 33 were kidnapped. Furthermore, 86 health facilities were damaged and 28 ambulances were targeted.

**Fig 1.**
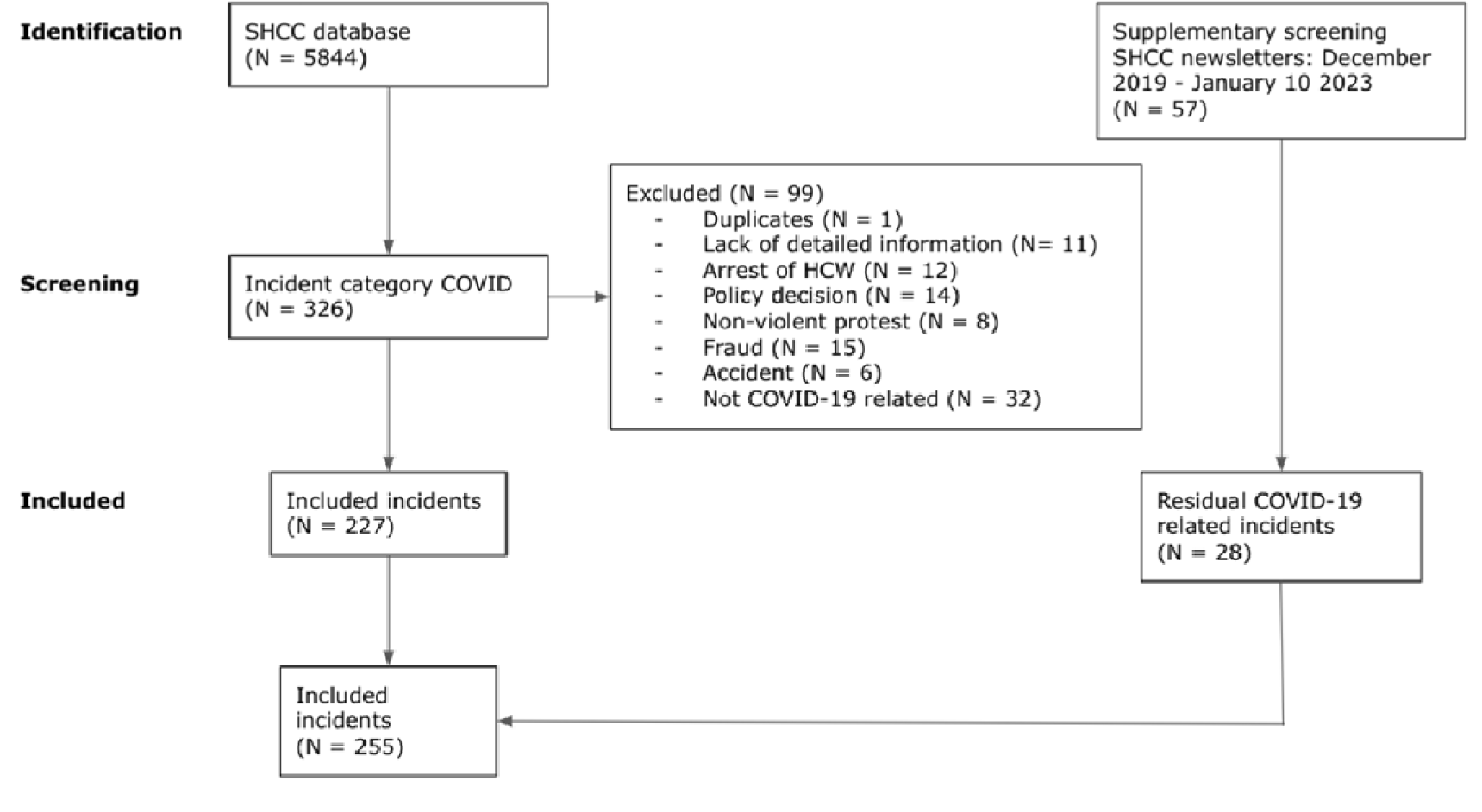
PRISMA flowchart. Abbreviations: SHCC = Safeguarding Health in Conflict Coalition, HCW = healthcare worker

### Attacks per geographic world region

Figure 2 depicts the incidence and impact of attacks against healthcare per world region. The world region with the highest number of reports was ‘Asia’ with 84 incidents (32.9%). ‘North America’ and ‘Middle East & North Africa’ were also frequently affected with 54 (21.1%) and 39 (15.3%) incidents, respectively. In Europe, facility attacks were predominant. In other world regions, incidents mostly involved direct attacks on HCWs. The countries with the highest numbers of reports were Mexico (n = 43; 16.9%), Myanmar (n = 33; 12.9%) and India (n = 30; 11.8%).

**Fig. 2.**
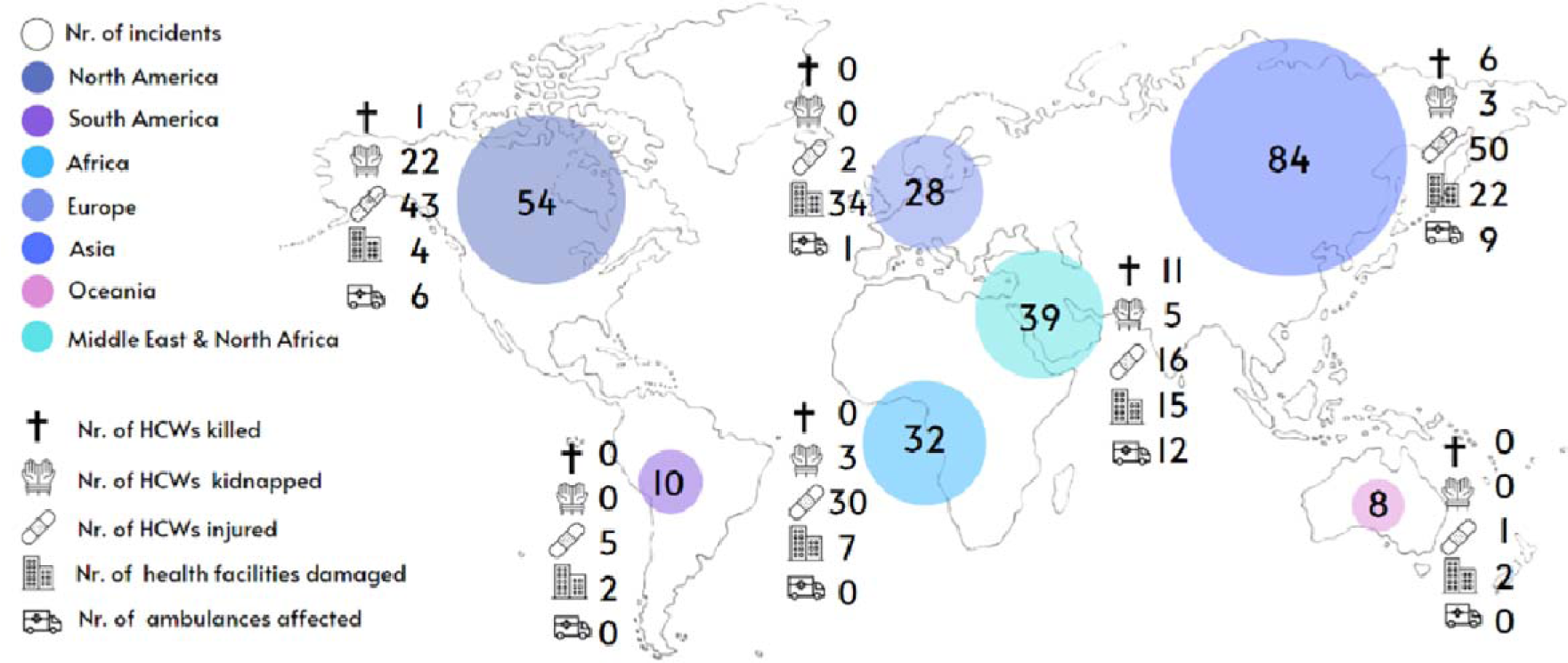
COVID-19 related attacks against healthcare per world region.* Abbreviations: HCWs = healthcare workers *Includes multiple incidents Chi squared tests showed a significantly different distribution of attack incidence (χ² = 116.62, P = <.001), killed HCWs (χ² = 43.47, P = <.001), kidnapped HCWs (χ² = 78.86, P = <.001), injured HCWs (χ² = 116.57, P = <.001), damaged health facilities (χ² = 71.72, P = <.001) and affected ambulances (χ² = 37.51, P = <.001).

**Fig. 3.**
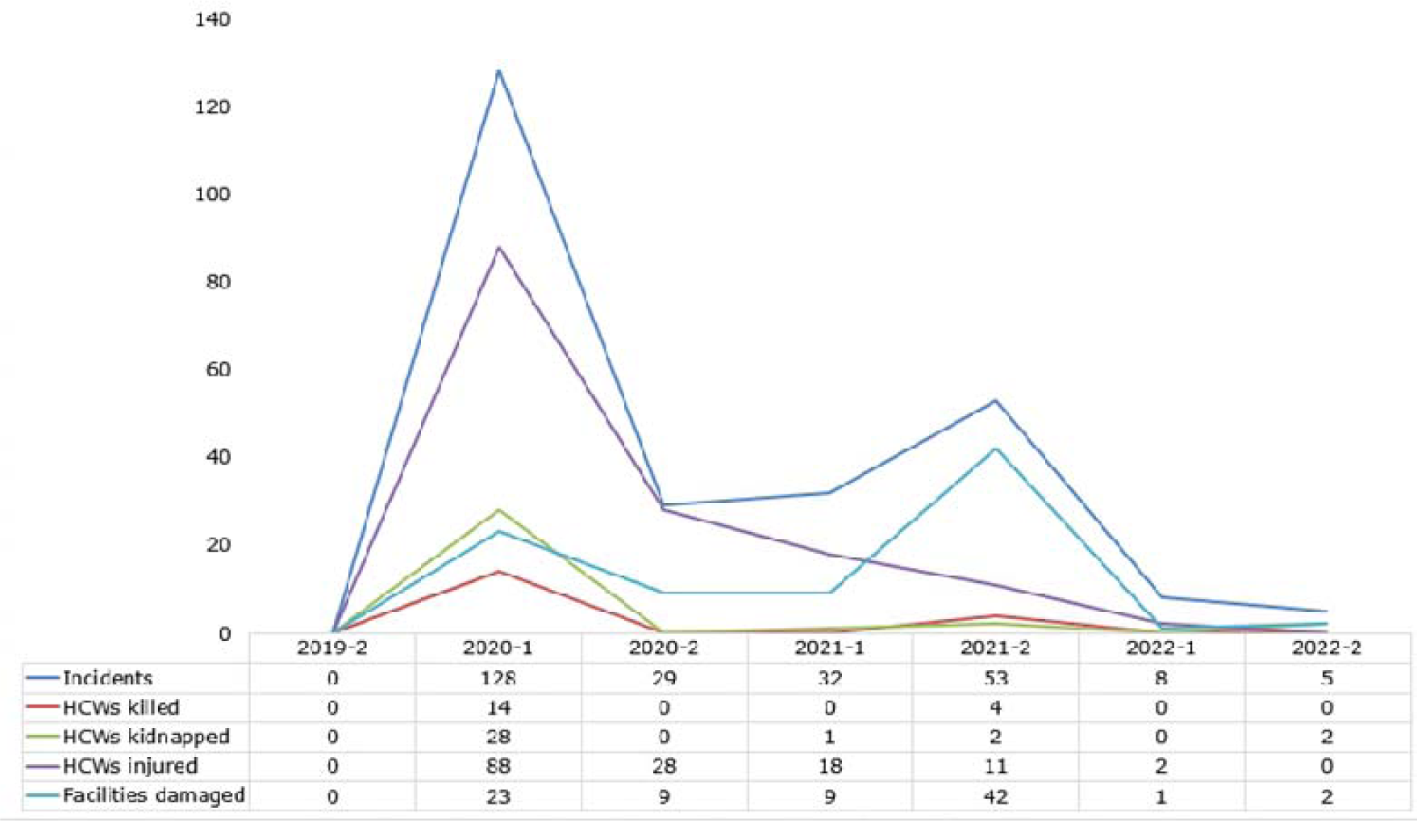
COVID-19 related attacks against healthcare per six-month time period.* Abbreviations: HCWs = healthcare workers *Includes multiple incidents Chi-squared tests showed a significant difference between incidence when comparing six-month time periods (χ² = 242.5, P = <.001)

### Incidence over time

Two periods with a peak incidence were observed. Most incidents were reported during the first peak in the first half of 2020 (n = 128; 50.2%). The second peak took place during the second half of 2021 (n = 53; 20.8%). The first peak was associated with the highest number of HCWs affected (killed, injured or kidnapped). During the second peak, facility attacks were the most predominant attack type.

### Temporal assessment of motives

The most commonly identified motives were conflict-related incidents targeting COVID-19 healthcare (n = 59; 23.1%), COVID-19 related stigma (n = 37; 14.5%), objection to medical health measures (n = 24; 9.4%), objection to vaccinations or the handling of vaccinations (n = 21; 8.2%), objection to public health measures (n = 16; 6.3%) and lockdown-associated police aggression directed at HCWs (n = 16; 6.3%). Incidents motivated by COVID-19 related stigma were most prevalent in the first half of 2020, whereas incidents associated with the large-scale vaccination campaign and conflict-related incidents were more frequent later in the course of the pandemic (Fig. 4). In the second half of 2021, all conflict-related incidents targeting COVID-19 healthcare were associated with the coup d’état in Myanmar (21 total). Other countries where conflict-related incidents targeting COVID-19 healthcare were repeatedly reported throughout the pandemic were the state of Palestine (n = 6), Yemen (n = 6) and Libya (n = 5). Less common motives listed were patient frustration, objection to buildings being re-allocated to treatment of COVID-19 patients, conspiracy theories, attacks targeting HCWs speaking out against difficulties in their work or criticizing COVID-19 policy and objection to scientific research regarding COVID-19. In 27 incidents, motives could not be derived from the incident description or context.

**Fig. 4.**
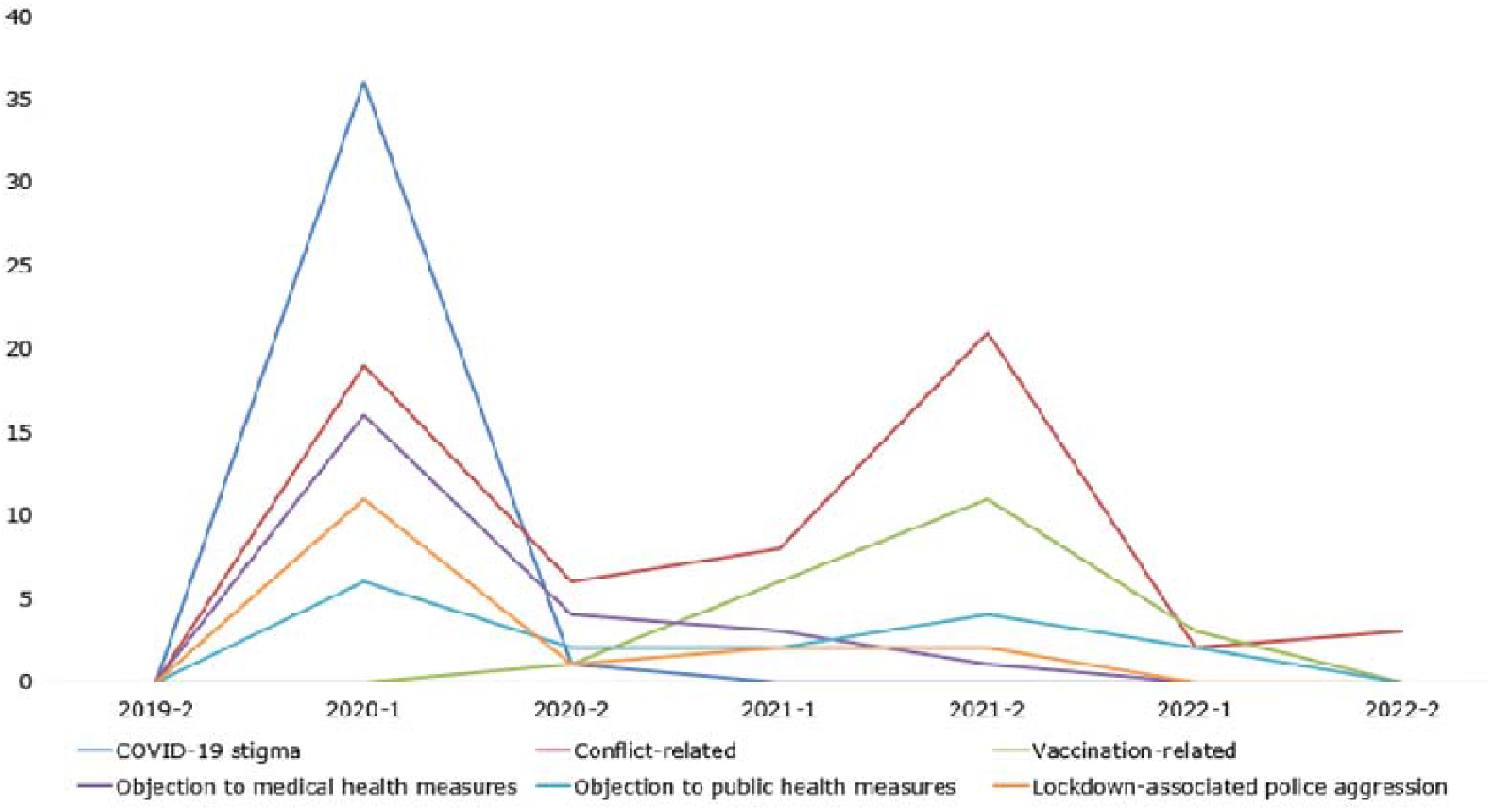
Most frequent motives over time.

**Fig. 5.**
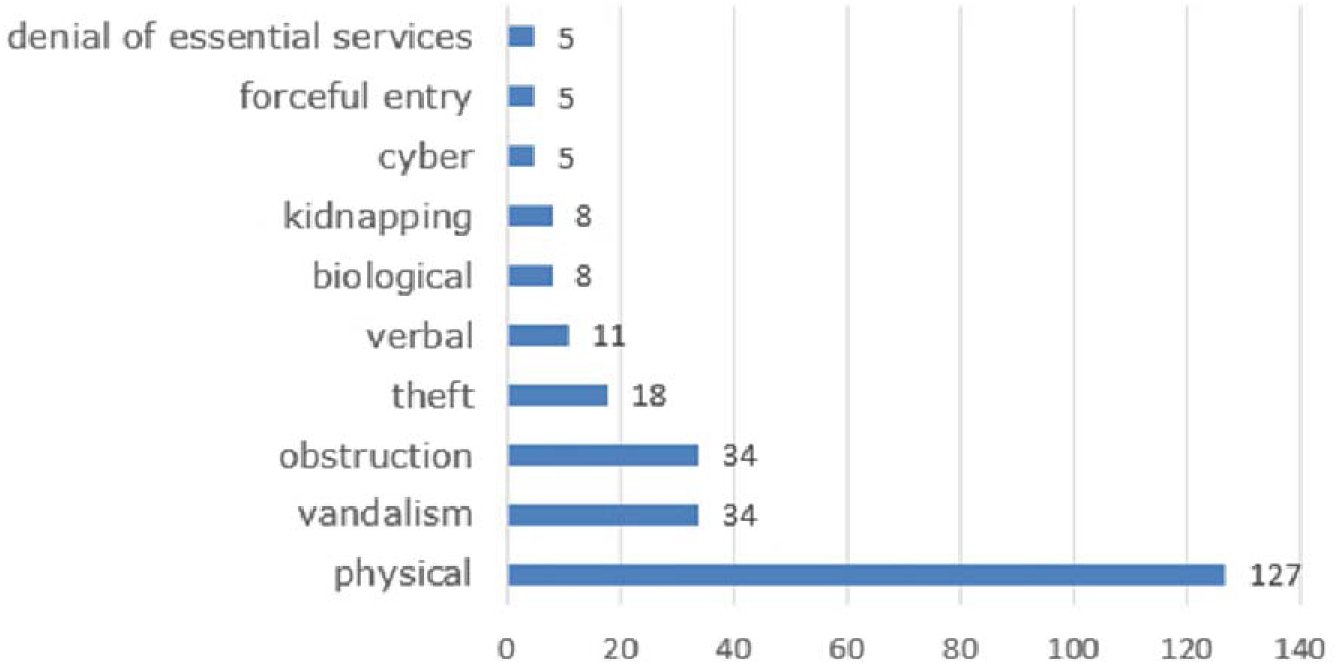
Types of COVID-19 related attacks on healthcare.

**Fig. 6.**
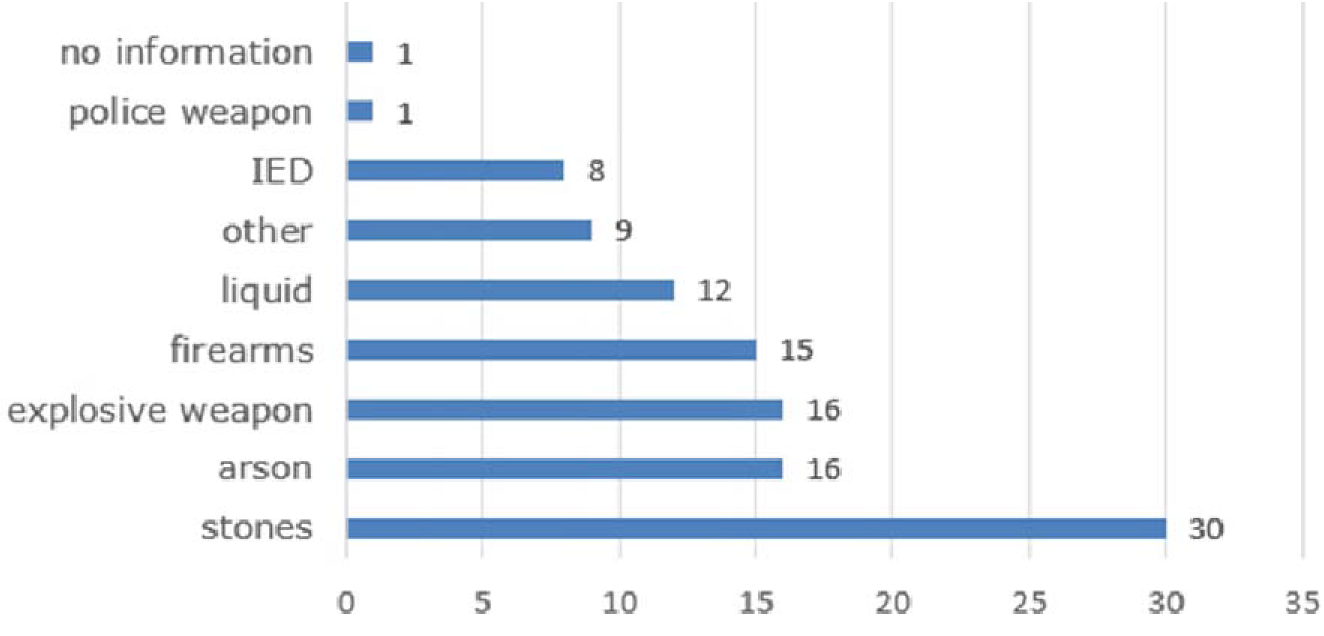
Weapons used in COVID-19 related attacks on healthcare. Abbreviations: IED = improvised explosive device

### Settings

Most incidents occurred in hospital settings (n = 39; 15.3%) (Table 1). The incidents which resulted in damaged or destroyed health facilities were mainly targeted against vaccination centers (28) and quarantine centers (18). Other common settings were “on the way to/from work” (n = 34; 13.3%) and “during community outreach” (n = 22; 8.6%).

**Table 1:**
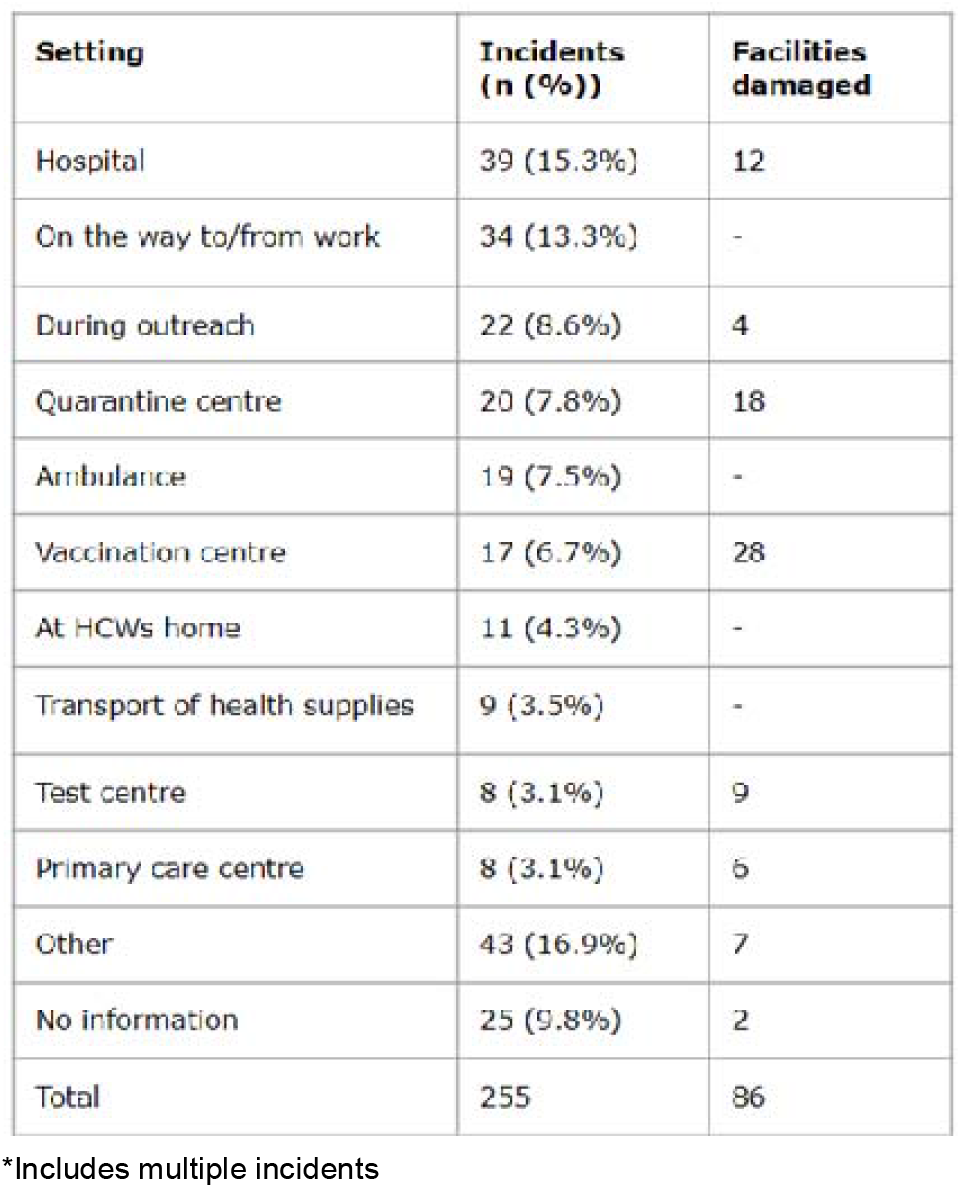
Settings where COVID-19 related attacks against healthcare occurred*.

**Table 2.** Incidents and impact on healthcare per perpetrator type*.

| Perpetrator<br>category | Incidents | HCWs killed | HCWs<br>kidnapped | HCWs<br>injured | Facilities<br>damaged | Ambulances<br>affected |
| --- | --- | --- | --- | --- | --- | --- |
| Employee | 6 (2.4%) | 0 | 0 | 0 | 1 (1.2%) | 0 |
| NSA | 14 (5.5%) | 3 (16.7%) | 5 (15.2%) | 4 (2.7%) | 8 (9.3%) | 9 (32.1%) |
| Patient/relative | 35 (13.7%) | 0 | 9 (27.3%) | 40 (27.2%) | 4 (4.7%) | 1 (3.6%) |
| Anti-government<br>extremist | 11 (4.3%) | 0 | 0 | 0 | 25 (29.1%) | 0 |
| State forces | 70 (27.5%) | 12 (66.7%) | 3 (9.1%) | 25 (17.0%) | 19 (22.1%) | 2 (7.1%) |
| Mob | 22 (8.6%) | 0 | 0 | 10 (6.8%) | 8 (9.3%) | 9 (32.1%) |
| Other | 29 (11.4%) | 1 (5.6%) | 3 (9.1%) | 26 (17.7%) | 1 (1.2%) | 1 (3.6%) |
| No information | 68 (26.7%) | 2 (11.1%) | 13 (39.4%) | 42 (28.6%) | 20 (23.3%) | 6 (21.4%) |

Abbreviations: NSA = non-state actor, HCWs = healthcare workers
\*Includes multiple incidents

### Attack types and weapon use

The most frequently identified attack type was physical violence (n = 127; 49.8%), followed by vandalism (n = 34; 13.3%) and obstruction (n = 34; 13.3%) (fig. 4). Physical violence was the most frequently identified type of violence (37.5% - 66.7%) in America, Africa and Asia (37.5% - 66.7%), whereas vandalism was most frequently reported in Europe and Oceania (25.0% −46.4%). Weapons were used in 106 (41.6%) incidents. The use of stones (n = 30; 11.8%) was most frequently reported, followed by arson (n = 16; 6.3%) and explosive weapons (n = 16; 6.3%).

### Perpetrators

A perpetrator type was identified in 73.3% of all cases. The most frequently reported perpetrator category concerned “state forces”, including military, police and policymakers (n = 70; 27.5%). In the Middle East & North Africa, Asia, and Africa, respectively 48.7%, 45.2% and 31.3% of incidents were attributed to state forces. State forces were also associated with the highest number of HCWs killed (n = 12; 27.5%). The perpetrator type “patient/relative” was most often linked with injured HCWs (n = 40; 27.2%). Sixty percent of these incidents were perpetrated by relatives of patients, and 40% by patients. Anti-government extremists were reported to have damaged the highest number of facilities (n = 25; 29.1%), but were not associated with HCWs being killed, kidnapped, or injured. Anti-government extremists were the most frequently identified perpetrator group in Europe, being responsible for 21.4% of all attacks. Ambulances were most frequently targeted by “non-state actors” (n = 9; 32.1%) and “mob” violence (n = 9; 32.1%).

## Discussion

COVID-19 related attacks against healthcare occurred globally and throughout the course of the pandemic. At least 18 HCWs were killed, 147 HCWs were injured and 86 facilities were damaged or destroyed due to COVID-19 related attacks. Incidents were heterogeneous with regards to motives, attack types and outcomes. Two periods with a peak incidence of reports were observed. The first half of 2020 was associated with the highest number of incidents, most of which were related to stigmatization of HCWs. The second peak in reporting occurred in the second half of 2021 and included incidents associated with the large-scale vaccination campaign and conflict violence targeting COVID-19 healthcare in Myanmar. Incidents related to stigmatization can be expected in the beginning of a pandemic involving a novel pathogen, when little is known about a disease. Stigmatization can be linked to fear of acquiring the disease and HCWs have an occupational exposure to COVID-19. In the community HCWs can be viewed as a source of infection, leading to the belief that they should be isolated from the community. (27) As a result, HCWs may experience stigmatization and bullying. (28, 29) The manner in which information is communicated via the media can either mitigate or exacerbate the fear of infection and thus stigmatization. (30) What is needed are public education campaigns clarifying that HCWs do not pose a risk to the public. (29)

In contrast with other world regions, where physical attacks against HCWs were most prevalent, Europe and Oceania most frequently witnessed attacks that involved vandalism and facility damage. Vandalism is generally seen as a characteristic of specific-issue terrorism, a term used to describe terrorism focused on a specific issue rather than more widespread change (31) For example, terrorist attacks against abortion clinics mostly involved arson attacks and facility attacks, with relatively low casualty numbers. (32) Wide-spread media attention is more important to this category of perpetrator than inflicting deaths and injuries. A certain number of attacks against vaccination-, test- and quarantine centers may be regarded as such. (10) However, attacks against healthcare facilities, and hospitals in particular, have the potential to disrupt services causing further damage. Therefore, pandemic preparedness planning should include consideration of security enhancements. Medical facility hardening strategies include securing and guarding points of entry, armed security guards and closed-circuit television (CCTV). (11)

This study also shows that a portion of the attacks on HCWs occur outside medical facilities, on the way to and from work, or during community outreach. A number of these incidents were caused by police aggression against HCWs during lockdowns. Medical organizations should communicate with security forces to raise awareness of the right of access to healthcare, especially in times of health crisis. (33) Authorities should facilitate safe transport to and from work if necessary. (1) Moreover, improved coordination between medical organizations and security forces about community outreach schedules could improve HCW safety during these endeavors. States and non-state actors have a responsibility to respect humanitarian and human rights law and protect HCWs. (1, 15)

COVID-19 misinformation and conspiracy theories may cause distrust in the healthcare system. (34) Although only a small percentage of incidents were directly motivated by conspiracy theories in this study, conspiracy theories may strengthen the resistance to public health measures and vaccination rollout, and subsequently lower the threshold for extremist groups or lone wolves to conduct attacks against healthcare. (35) Community trust in healthcare is a necessary element to successfully implement public and medical health measures and to reach sufficient vaccination coverage. (36) Therefore, authorities should focus on engaging the public and providing reliable information throughout any pandemic.

The SHCC database relies on incident reports by organizations and media. However, not all violence is reported. Therefore, the number of attacks against healthcare reported is likely underestimated. In two meta-analyses, the pooled global prevalence of workplace violence against HCWs during the COVID-19 pandemic was found to be 43 - 47%, including physical and psychological violence. (37, 38) Frequently, HCWs who experienced violence do not report such incidents to anyone, and only a minority of cases are reported to the police. (37, 38) Reasons for not reporting are beliefs that reporting is useless, considering the incident as unimportant, and fear of negative consequences. (39, 40) However, as a result of under-reporting, violence against healthcare remains covert. Unreported incidents cannot be prosecuted and impunity for these violations remains.

Policy attention to attacks against healthcare has long been stuck in a familiar pattern: rapid condemnation of the attack, followed by inaction until another attack occurs. (15) In order to facilitate accountability, monitoring of attacks is a crucial first step that could be improved by addressing methodological challenges and political and institutional obstacles in public databases. (18) Moreover, reporting could be improved by a change of attitude towards violence against healthcare and amelioration of the legal system. This will also allow for more reliable research, which will help to determine efficacy and identify policy interventions.

The COVID-19 pandemic exposed the paradox HCWs faced: being applauded for their work, but at the same time silenced, stigmatized and attacked. Behind every statistic in this study, there are stories of HCWs and their patients suffering, which cannot be captured in numbers. To express outrage about these attacks is a first step, but not enough. The protection of healthcare should be prioritized in order to guarantee an effective public health response to future pandemics.

### Strengths and limitations

As this research is based on the SHCC database, it is limited by its design. The SHCC database is the most detailed and transparent database of worldwide attacks against healthcare, but relies on media reporting and is therefore subjected to information and selection bias. It can be inferred that the data presented are an underrepresentation, as many incidents go unreported. As such, the incidents collected are a minimum estimate of the actual magnitude of violence against healthcare during the COVID-19 pandemic. The data were gathered from open sources and SHCC contributors. Accuracy and reliability of the reported information are highly dependent on the source. Moreover, the number of contributors as well as the level of press freedom and human rights awareness differ among countries. The database collects most information from sources in English or French, leading to an underrepresentation of incidents in countries where those languages are not practiced. The data is likely to present a skewed view on attack types, because the more severe or more remarkable attacks are more likely to be reported. (22) Consequently, this also suggests that the incidents with a high impact are likely included.

A strength of this study is the level of detail and transparency in the analysis. Multiple categories were added to elaborate data extraction and a clear list of categories and definitions was provided. Additional searches were performed in multiple languages to provide more detailed information. Finally, this study is the first to analyze COVID-19 related attacks throughout multiple phases of the pandemic in the years 2020 - 2022.

## Conclusion

COVID-19 related attacks against healthcare occurred globally and throughout the course of the pandemic. There were two periods with a peak incidence of reports. The first peak occurred during the beginning of the pandemic, and mainly concerned stigma-related attacks against healthcare. The second peak, in 2021, was attributed to conflict in Myanmar and the global vaccination campaign. Government officials and public health officers should include consideration of context-specific protective measures for healthcare workers and facilities, especially during times of turmoil and during pandemics.

## Data Availability

https://map.insecurityinsight.org/health

## Acknowledgements

The authors would like to thank Insecurity Insight and the Safeguarding Health in Conflict Coalition for providing the data.

## Authorship contribution statement

W. Duffhues and D.Barten were responsible for conceptualization, incident selection, data extraction, data analysis and drafting of the manuscript. F. Van Osch supervised data analysis and drafting of the manuscript. H. De Cauwer, L. Mortelmans, M. Koopmans, D. Tin and G. Ciottone provided feedback and contributed to the final version of the manuscript. All authors read and approved the final manuscript.

## Authors’ disclosure

The authors declare that they have no known competing financial interests or personal relationships that could have appeared to influence the work reported in this paper.

## Funding statement

This research received no specific grant from any funding agency in the public, commercial, or not-for-profit sectors.

## Appendix 1 Categories and definitions

Healthcare workers killed: HCW’s that died due to violence. (Not: HCWs that have died from insufficient healthcare). In case the number is unknown this is noted.

Healthcare workers injured: HCW’s that are reportedly injured or reportedly physically attacked or assaulted. In case the number is unknown, this is noted.

Healthcare workers kidnapped: HCW’s that are reportedly kidnapped. Also includes virtual kidnappings (extortion schemes in which the perpetrators claim to have kidnapped a loved one). Does not include detention or arrest of HCW’s. In case the number is unknown, this is noted.

Facilities damaged: Healthcare facilities that are reportedly damaged. Includes facilities that are vandalized, that are attacked with explosives, firearms or arson. Also includes facilities that were raided or if a theft occurred that impaired functioning of the hospital and includes the obstruction of electricity to the hospital. In case the number is unknown, this is noted.

Ambulances targeted: includes ambulances that are reported damaged or stolen. In case the number is unknown, this is noted. Does not include obstruction of ambulance services.

### Settings

- Hospital: A health facility permanently staffed by at least one physician, which can offer inpatient accommodation and can provide active medical and nursing care. Hospitals may be classified by type of service, ownership, size by number of beds, and length of stay. It is a reservoir of critical resources and knowledge and provides a setting for education and research. (1)
- Vaccination center: A building specifically dedicated to (mass) vaccination against COVID-19.
- Test center: A building specifically dedicated to (mass) testing for COVID-19
- Quarantine center: A facility specifically designated for isolation and treatment of persons (possibly) infected with COVID-19. The facility is reported as a quarantine center or COVID-19 treatment center.
- Primary care center: A community-based clinic that provides the first level of basic or general healthcare for an individual’s health needs, including diagnostic and treatment services and, if integrated into the clinic’s service array, mental health services.
- Ambulance: A vehicle equipped for taking sick or injured people to and from hospital, especially in emergencies. The ambulance may also be used for transport of health supplies or for transport of the deceased.
- During outreach: The activity of providing services to any population that might not otherwise have access to those services. The health workers are mobile, it involves meeting someone in need of an outreach service at the location where they are. It includes the screening of people for COVID-19 in their neighborhood as well as COVID-19 checkpoints.
- Healthcare worker’s home: The place where a health worker is residing.
- On the way to/from work: The health worker was en route to/from work when the incident happened. (2)
- Transport of health supplies: the attack was on a transport of health supplies regardless of the transport vehicle. Does not include attacks on ambulances if used for transporting supplies.
- Other: Any other place that is not a health facility, ambulance, during outreach or traveling to/from work or the health worker’s home.
- No information

Note: the setting corresponds to the function of the building at the time of the incident. Eg. when an Ebola clinic was used for COVID-19 quarantine, the setting was categorized as ‘quarantine center’ instead of ‘other’.

### Attack types

- Verbal: The use of words or gestures to cause psychological harm. It involves threatening, screaming at, or cursing people. It includes attacks perpetrated per phone but not online attacks.
- Physical: Aggression that involves harming others physically—for instance hitting, kicking, stabbing, or shooting them; aggression that involves damaging or destructing healthcare materials and/or materials.
- Cyber: Any of the following: any attempt to gain unauthorized access to a computer, computing system or computer network with the intent to cause damage; online threats or harassment.
- Obstruction: Restricting an ambulance or other medical transport access to a region; prohibiting health workers from providing care or traveling to work; blocking entry and/or exit to a health facility or restricting access to care. It also includes arrests of healthcare workers clearly violating human rights.
- Forceful entry: The unlawful taking of possession of real property by force or threats of force or unlawful entry into or onto another’s property, especially when accompanied by force.
- Theft: The stealing of medical materials and/or the plundering of a health facility or ambulances. Also includes stealing bodies of the deceased.
- Biological: Includes spitting or coughing on a healthcare worker.
- Kidnapping: Includes taking a health worker against their will. Also includes virtual kidnappings. Does not include detention or arrest of HCW’s.
- Denial of essential services: denial of services essential for HCW’s to do their job or for healthcare facilities to function, for example eviction from home of denial of public transport to/from work
- Vandalism: the intentional damaging of property. Does not include incidents in which persons were attacked as well.
- No information

Note: in case of overlapping attack types the primary attack type was noted. Attack types other than verbal or physical do not exclude that any physical or verbal violence was used.

### Weapons used

- Yes: weapons were used. This may also include materials not specifically designed as a weapon.
- No: no weapons were used. Incidents are also put into this category if weapons were carried by the perpetrator but not used.
- No information

### Weapon types

- Arson: Any instrument or material associated with fire-making (e.g. matches, kerosene, propellant). (2)
- Firearms: Any of the following: assault rifle (e.g. Kalashnikov AK47 or variant, M16, etc.), machine gun, submachine gun, pistol, or revolver. (2)
- Explosive weapon: Any of the following: aerial bomb, missile, mortar shell. (2)
- IEDs (improvised explosive device): Any of the following: car, suicide or roadside bomb improvised from materiel not specifically designed for that purpose; grenade. (2)
- Knife: Any of the following: ax, blunt instrument, dagger, machete or sword. (2)
- Liquid: Any of the following: anti-bacterial gel, bleach, hot beverage or cooked rice/gruel. (2)
- Police weapon: Any of the following: taser, live or rubber bullet, tear gas or water cannon. (2)
- Stones: Any of the following: rocks, stones, sticks. (2)
- Other: the weapon type used is reported but does not fit in any of the named categories.
- No information

### Perpetrators

- Employee: A current or former employee of the aid agency or health provider. (2)
- NSA (non-state actor): A perpetrator who is part of a named or unnamed armed group that is not part of the state’s law enforcement, military, or security apparatus and who is either involved in a state-based armed conflict with a government or is engaged with an NSA in a non-state conflict, or engages in one-sided violence, based on Uppsala Conflict Data Program definitions. This includes all organized armed groups such as private armies, rebel or guerrilla groups, and terrorist groups. It includes unidentified or unnamed groups of armed individuals (e.g. a group of unidentified ‘armed men’) if the incident description refers in a generic sense to rebels or extremists or groups in some form affiliated with the military or using military-type structures or equipment (e.g. wearing army fatigues, etc.), without indicating a linkage to any state army. This does not include private security actors or unidentified or unnamed groups of armed individuals (e.g. a group of unidentified ‘armed men’) who commit robberies, burglaries, theft or fraud. (2)
- Patient/relative: The direct beneficiary of healthcare or a relative/caregiver of a patient receiving healthcare.
- State forces: Includes police, military and/or policymakers.
- Mob: a large and disorderly crowd of people. The perpetrators are described as a mob or a group of rioters.
- Anti-government activists: A person that primarily opposes the government, both issue-driven and ideological. They object to governments that recognize the COVID-19 pandemic and take measures to mitigate it. The government is seen as the primary enemy. The perpetrator(s) should have expressed these views. (3,4)
- Other: the perpetrator is a known actor but does not fit into any of the other categories
- No information

Note: the perpetrator category corresponds to the role of the perpetrator at the time the incident took place. Eg. in case a perpetrator was ‘NSA’ at the time of the attack, but later became ‘state forces’, the perpetrator is categorized as NSA.

### Motives

- Objection to hospitals or buildings being used to treat COVID-19 patients: Resistance to hospitalization to treat COVID-19 in a hospital ward; opposition to the use of hospitals to treat COVID-19 patients; opposition to the presence of quarantine centers. (2)
- Objection to medical health measures: Resistance to medical tests to check for COVID-19 infections or antibodies; opposition to changes in local burial or cremation customs and practices, such as prohibiting relatives from attending funeral ceremonies, or from seeing the deceased before burial/ cremation; and protests against the use of specific burial locations for COVID-19 victims.(2)
- Objection to public health measures: Resistance to disinfection; attacks on workers carrying out disinfection activities; resistance to COVID-19 sensitization campaigns; attacks on individuals engaged in sensitization campaigns; opposition to social distancing measures that take the form of protests or violence against health providers; or resistance to contact-tracing activities. (2) Also includes opposition to wearing a mask.
- Response to whistle blowing: Retaliation against or silencing of HCWs speaking out against difficulties in their work, such as the lack of personal protective equipment or being criticized/disciplined for reporting higher infection rates than those of the government. (2) Also includes speaking out against the government’s COVID-19 policy. It does not include HCWs asking people to adhere to COVID-19 medical or public health measures.
- Objection to (the handling of) vaccinations: Resistance or opposition to COVID-19 vaccinations or other vaccination campaigns; or protests related to the mismanagement of vaccination supplies. (2) Also includes theft or destruction of vaccines.
- Conflict-related: An incident is categorized as conflict-related when the perpetrator(s) are listed as a conflict party in the Uppsala Conflict Data Program (UCDP). UCDP uses the threshold of 25 fatalities per calendar year attributable to a single perpetrator or during confrontations between two perpetrator groups before a perpetrator is added to the list. (2)
- Research-related: Resistance to COVID-19 related research. Does not include contact-tracing activities.
- Stigmatization: acts driven by social and physical avoidance (e.g. denial of bus service or eviction from home) and acts expressing disgust (5), such as spitting or coughing on a HCW or an attack with bleach. Also includes attacks in which the perpetrator accuses HCWs of spreading the virus.
- Law enforcement aggression during lockdown: State forces obstructing or assaulting HCW’s en route to/from work during a lockdown; State forces obstructing or assaulting ambulances during a lockdown.
- Patient frustration: violence perpetrated by patients or caregivers triggered by healthcare not corresponding to patient expectations, for example treatment not being available, death of a covid-19 patient or otherwise perceived insufficient care.
- Conspiracy theory: the perpetrators’ actions were motivated by conspiracy theories, e.g. government using coronavirus to make people sick, or coronavirus being used for political or financial gain.
- No information

Note: in case of overlapping motives the primary motive was noted.

### Sources

1. WHO. Hospitals. WHO website. Accessed December 19, 2022. https://www.who.int/health-topics/hospitals#tab=tab_1
2. Insecurity Insight. Attacked and Threatened: Healthcare at risk Categories and definitions 2021. Insecurity Insight website. Published May 2021. Accessed December 17, 2022. https://insecurityinsight.org/wp-content/uploads/2020/11/Categories-and-definitions-Attacked-and-Threatened-Health-care-targeted-in-conflict-and-COVID-19.pdf.
3. Jackson S. What is Anti-Government Extremism. Perspectives on Terrorism. 2022;16(6):9-18.
4. Van Dongen T. Assessing the Threat of Covid 19-Related Extremism in the West. ICCT website. Published August 5, 2021. Accessed December 19, 2022. https://icct.nl/publication/assessing-the-threat-of-covid-19-related-extremism-in-the-west-2/.
5. Oaten M, Stevenson RJ, Case TI. Disease avoidance as a functional basis for stigmatization. Philos Trans R Soc Lond B Biol Sci. 2011;366(1583):3433-52.

## Notes

### Competing Interest Statement

The authors have declared no competing interest.

### Funding Statement

This study did not receive any funding

### Author Declarations

This was a review of the Safeguarding Health in Conflict Coalition Database, which is openly available to anyone. Source: https://map.insecurityinsight.org/health

